# Detection and Characterization of Thrombosis in Humans using Fibrin-Targeted Positron Emission Tomography and Magnetic Resonance

**DOI:** 10.1101/2020.08.17.20176172

**Authors:** David Izquierdo-Garcia, Pauline Désogère, Anne L. Philip, Choukri Mekkaoui, Rory B. Weiner, Onofrio A. Catalano, Yin-Ching Iris Chen, Doreen DeFaria Yeh, Moussa Mansour, Ciprian Catana, Peter Caravan, David E. Sosnovik

## Abstract

The detection of thrombus in the left atrial appendage (LAA) is vital in the prevention of stroke. We present a novel technique to detect and characterize LAA thrombus in humans using positron emission tomography (PET) of a fibrin-binding radiotracer, [^64^Cu]FBP8. Initial testing in healthy volunteers (n = 8) revealed that [^64^Cu]FBP8 was stable to metabolism and was rapidly eliminated with a blood half-life of one hour. Patients with atrial fibrillation (AF) and recent transesophageal echocardiograms (TEEs) of the LAA (positive n = 12, negative n = 12) were studied. PET, integrated with magnetic resonance (PET-MR), of the thorax was performed one hour after [^64^Cu]FBP8 injection. The maximum standardized uptake value (SUV_Max_) in the LAA was significantly higher in the TEE positive than negative subjects, median [interquartile range] of 4.0 [3.0–6.0] vs. 2.3 [2.1–2.5]; p < 0.001. A SUV_Max_ threshold of 2.6 correctly identified 12/12 positive TEEs and 10/12 negative ones, yielding an area-under-the-receiver operating characteristic curve of 0.97. The minimum longitudinal magnetic relaxation time (T1_Min_) in the LAA was significantly shorter in the TEE positive than TEE negative group 970 [780–1080] vs. 1380 [1120–1620], p < 0.05, with some overlap between the groups. Logistic regression using SUV_Max_ and T1_Min_ allowed all TEE positive and negative subjects to be classified with 100% accuracy. A strong correlation was seen between fibrin (SUV_Max_) and methemoglobin (T1_Min_) content in the LAA. In conclusion, the detection of LAA thrombus in humans with PET-MR of [^64^Cu]FBP8 is highly accurate and provides useful information on the biological properties of cardiac thrombus.

**One Sentence Summary:** First-in-human fibrin-targeted PET-MR of thrombus

## Introduction

Thrombosis plays a central role in numerous cardiovascular diseases and is associated with high morbidity and mortality (*1*). While major advances have been made in the molecular understanding and treatment of thrombosis, its diagnosis remains indirect and imprecise *(2, 3)*. The presence of a filling defect on computed tomography (CT) and magnetic resonance imaging (MRI), or the detection of echogenic material on ultrasound, currently form the basis of non-invasive thrombus detection. These approaches are of major clinical utility but provide no information on the molecular and cellular properties of the thrombus. In addition, these techniques frequently employ nephrotoxic contrast agents, have limited anatomical coverage and, in the case of transesophageal echo (TEE), involve heavy sedation and the risk of esophageal trauma.

Molecular imaging involves the use of imaging agents that are targeted to a specific molecular moiety and holds the promise of improving diagnostic accuracy and providing specific biological information on the target of interest. In the case of thrombosis this would include thrombus age, platelet content, fibrin content, risk of embolism and potential for lysis. A number of molecular imaging approaches incorporating different imaging modalities and targeting platelets (*4–10*), fibrin (*11–16*), d-dimers (*17–20*), or fibrinogen (*21*), have been proposed over the years, with some progressing to clinical trials (*3, 22*). However these agents have been limited by suboptimal pharmacokinetics, high blood background, low specificity, or sensitivity only to hyperacute/acute thrombi(*3*). The advantages of targeting fibrin for thrombus detection have been long recognized with initial (*13, 14*), and more recent (*23, 24*), descriptions of this approach using antibodies or antibody fragments against fibrin. We have previously described the use of a small fibrin-avid peptide to image thrombosis by MRI (*11, 25, 26*). The translational potential of this agent, however, was limited motivating the design of a new agent with full translational potential.

We have recently reported the preclinical development of a small peptide based fibrin-specific probe labeled with the copper-64 isotope to detect thrombus by positron emission tomography (PET) (*27*). This probe, [^64^Cu]FBP8, has submicromolar affinity for fibrin, and facilitated the rapid detection of venous, arterial, and embolic thrombi in a number of animal models (*27–30*). Here, we report the first human experience with [^64^Cu]FBP8, which we use to detect left atrial appendage (LAA) thrombus in subjects with atrial fibrillation. The detection of LAA thrombus was selected due to the clinical prevalence of atrial fibrillation and stroke (*31*), because TEE provides an accurate gold standard for comparison (*32*), and because the use of LAA occlusion devices in selected individuals allows the development of the thrombus to be precisely timed (*33–35*). We further aimed in the study to assess whether an integrated PET-MR approach, enabling the simultaneous acquisition of PET and longitudinal magnetic relaxation (T1) maps of the heart would enhance diagnostic accuracy and help characterize the detected thrombi. This study is, to the best of our knowledge, the first description of thrombus-targeted imaging in humans using integrated PET-MR.

## Results

### Safety and Pharmacokinetics of [^64^Cu]FBP8 in Healthy Human Volunteers

To determine the safety and pharmacokinetics of [^64^Cu]FBP8 in humans, initial investigation of the probe was performed in 8 healthy volunteers. The healthy controls (HC) were injected, while in the scanner, with an average [^64^Cu]FBP8 dose of 380 ± 140 MBq and imaged at 5-minute intervals for 2 hours, and then at 4–5 and 18–36 hours post injection. In addition, venous blood samples were drawn at multiple time points post-injection.

The structure of [^64^Cu]FBP8 is provided in Figure 1A. The probe is prepared by heating the peptide-chelator precursor with [^64^Cu]CuCl_2_ in sodium acetate buffer at 60 °C for 40 min. No further purification is required. After intravenous administration, the relatively low molecular weight of 2300 Da facilitates rapid and efficient excretion into the urine (Figure 1B). A smaller fraction of the radioactivity is excreted via the hepatobiliary system. Time activity curves (TACs) from regions of interest (ROIs) drawn in the left ventricle (blood pool), lung, and background (measured in a large ROI capturing the gluteus maximus and/or the quadriceps muscles) show that [^64^Cu]FBP8 is rapidly cleared from these tissues (Figure 1B), with little activity remaining by 4 hours. High performance liquid chromatography (HPLC) of the blood samples revealed that the majority of ^64^Cu activity was present as [^64^Cu]FBP8, which has a retention time of 12 minutes (Figure 1C). While some Cu-64 containing metabolites were present, approximately 50% of the circulating radioactivity was still due to intact [^64^Cu]FBP8 two hours post injection (Figure 1D). In addition, the probe remained biologically active in the blood and a substantial fraction of circulating Cu-64 activity retained the ability to bind fibrin (Figure 1E), even 2 hours post-injection. Radioactive copper was eliminated from the blood in a mono-exponential fashion with a blood half-life of 56.7 ± 6.6 min (Figure 1F), closely resembling the TAC in the blood. A visual depiction of [^64^Cu]FBP8 biodistribution is provided in Figure 1G, and a magnified view of the heart in Figure 1H. Note that the standardized uptake value (SUV) scale used in the magnified image (Figure 1H) is narrower. The background signal in the heart and thorax was low within 1 hour of injection and was virtually eliminated within 4 hours of injection.

**Figure 1.**
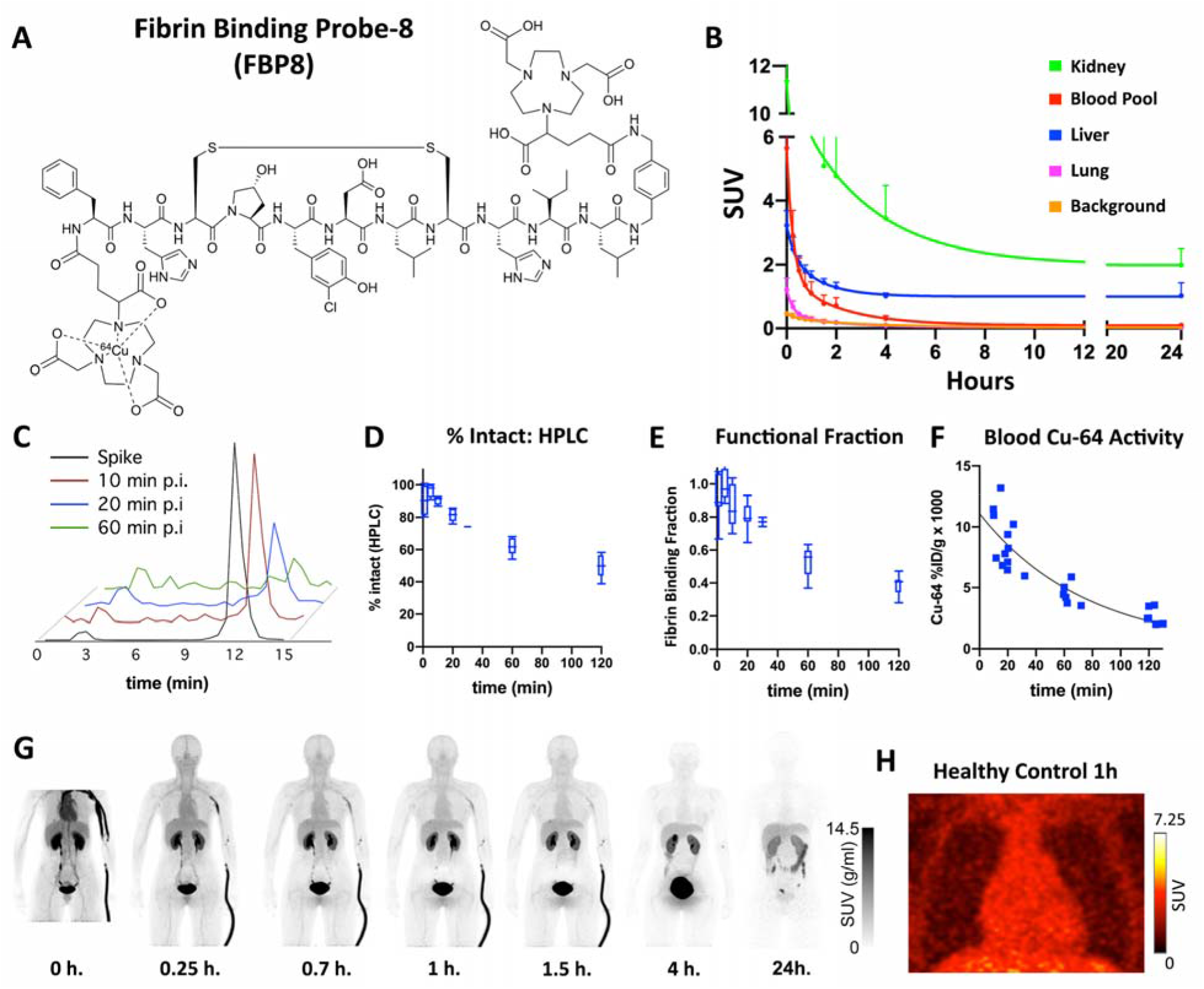
Structure and properties of ^64^Cu-FBP8. (A) The probe consists of a 6 amino-acid cyclic peptide with a high affinity for fibrin, conjugated to a ^64^Cu-containing chelator (NODAGA). (B) Average time activity curves (TACs) from 5 healthy subjects, expressed as SUV (g/ml), follow a biexponential model with a relatively rapid reduction of the PET signal in the blood and lung, accompanied by renal and hepatic elimination. (C-F) Chemical analysis of blood samples taken from healthy volunteers injected with [^64^Cu]FBP8. (C) HPLC of ^64^Cu-containing moieties in the blood at various times p.i. shows that the majority of ^64^Cu activity is intact [^64^Cu]FBP8 which has a retention time of 12 minutes. (D) [^64^Cu]FBP8 is stable in blood, with >60% of the probe still intact by HPLC one hour after injection. (E) Over 50% of the probe remains functional and able to bind fibrin 1 hour after injection. (F) The clearance of ^64^Cu activity from the blood, expressed as %ID/g, indicates a one-hour blood half-life. (G-H) Biodistribution of [^64^Cu]FBP8 in a healthy control subject shows a substantial reduction in the background thoracic signal within 1 hour of injection, and a near-complete loss of background by 4 hours. Elimination of the probe in the bladder and gallbladder is clearly seen. (H) Magnified view of the thorax at 1 hour showing no evidence of focal [^64^Cu]FBP8 uptake in either the heart or lungs.

Serological testing in the healthy controls revealed no changes in renal or hepatic function, no changes in the complete blood count and no changes in markers of coagulation. Probe injection did not result in the prolongation of the QT interval and did not produce any clinical symptoms in any of the study participants.

### Comparison of [^64^Cu]FBP8 versus TEE in Subjects with Atrial Fibrillation

We next examined the ability of [^64^Cu]FBP8 PET to detect LAA thrombus in patients (n = 24) with atrial fibrillation (AF) and a recent TEE. The AF cohort consisted of subjects with no evidence of LAA thrombus by TEE (n = 12) and subjects with confirmed LAA thrombi (n = 12). The TEE positive cohort consisted of patients with spontaneous LAA thrombi (n = 4) and those with LAA thrombi purposefully induced through the placement of a LAA occlusion device (Watchman, Boston Scientific). The age of the spontaneous thrombi detected by TEE could not be determined, but their size measured by TEE ranged from 7 – 14 mm in two dimensions (mean 2D area 93 mm^2^). The induced LAA thrombus behind the Watchman device, in subjects who received it, was assumed to occur on the day of implantation and thrombus age in this population was 5 days (n = 3), 12 days (n = 4) and 54 days (n = 1). Subjects with chronic, organized thrombi were not included in the study. Subjects who were on anticoagulation therapy prior to their TEE were included in the study, but were excluded if intravenous or oral anticoagulation was initiated after their TEE. TEEs with spontaneous echo contrast were classified as negative by the clinical caregivers and formed part of the negative TEE cohort in this study. The characteristics of the subjects in the TEE positive and negative cohorts are described in the Table 1. The majority (21/24) of subjects were taking direct oral anticoagulants (DOACs) at the time of imaging, 2 were taking warfarin and 1 was not taking any anticoagulant. Five of the subjects in the TEE positive group, and one in the TEE negative group, had a history of TIA, stroke or thromboembolism. One of the subjects in the study (in the TEE negative group) had a history of a hypercoagulable state (Factor V Leiden). All recruited subjects were imaged with [^64^Cu]FBP8 PET within 2 weeks of their TEE. PET-MR imaging with [^64^Cu]FBP8 was performed 1 – 2 hours after intravenous injection of the probe.

**Table 1:**
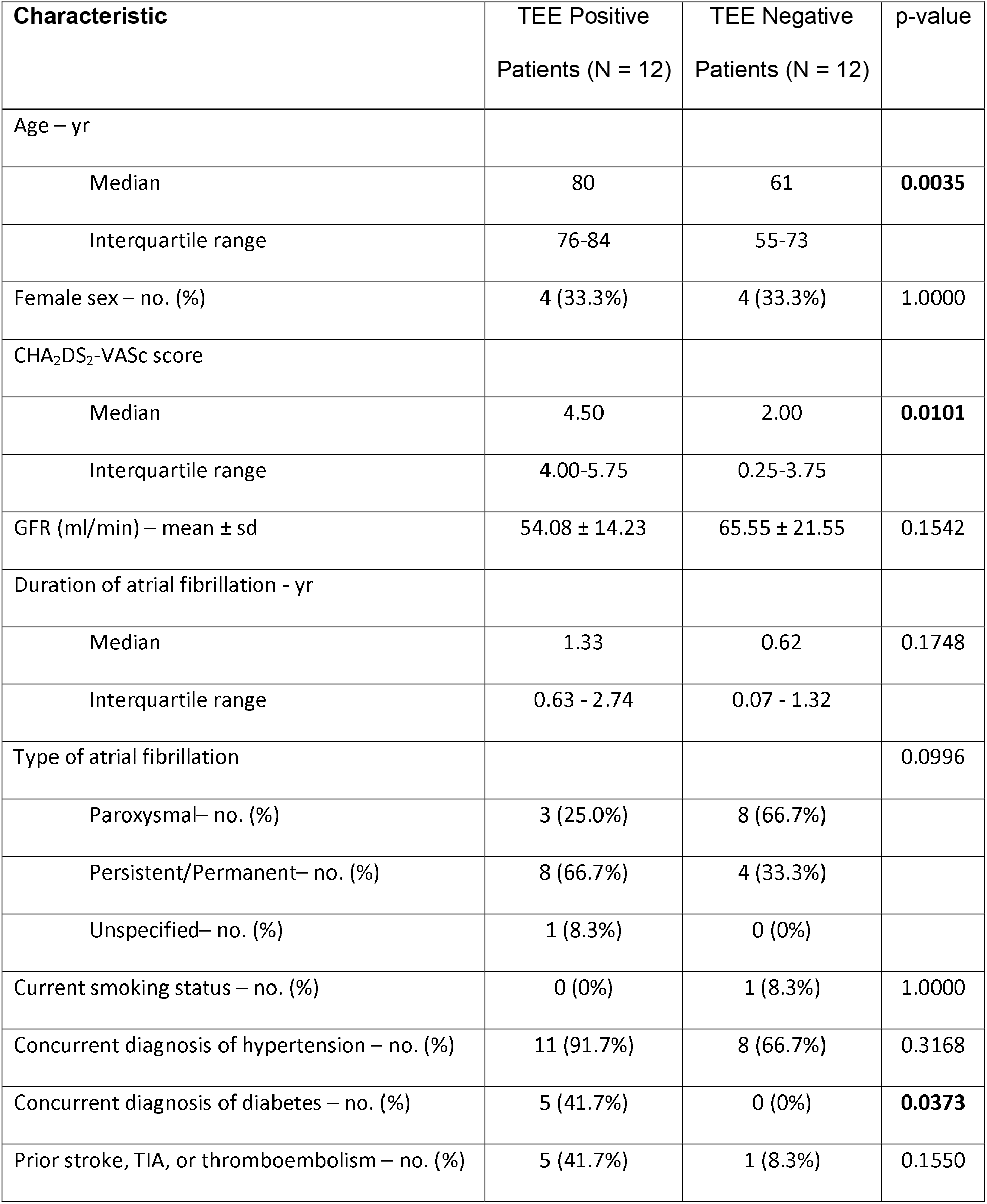
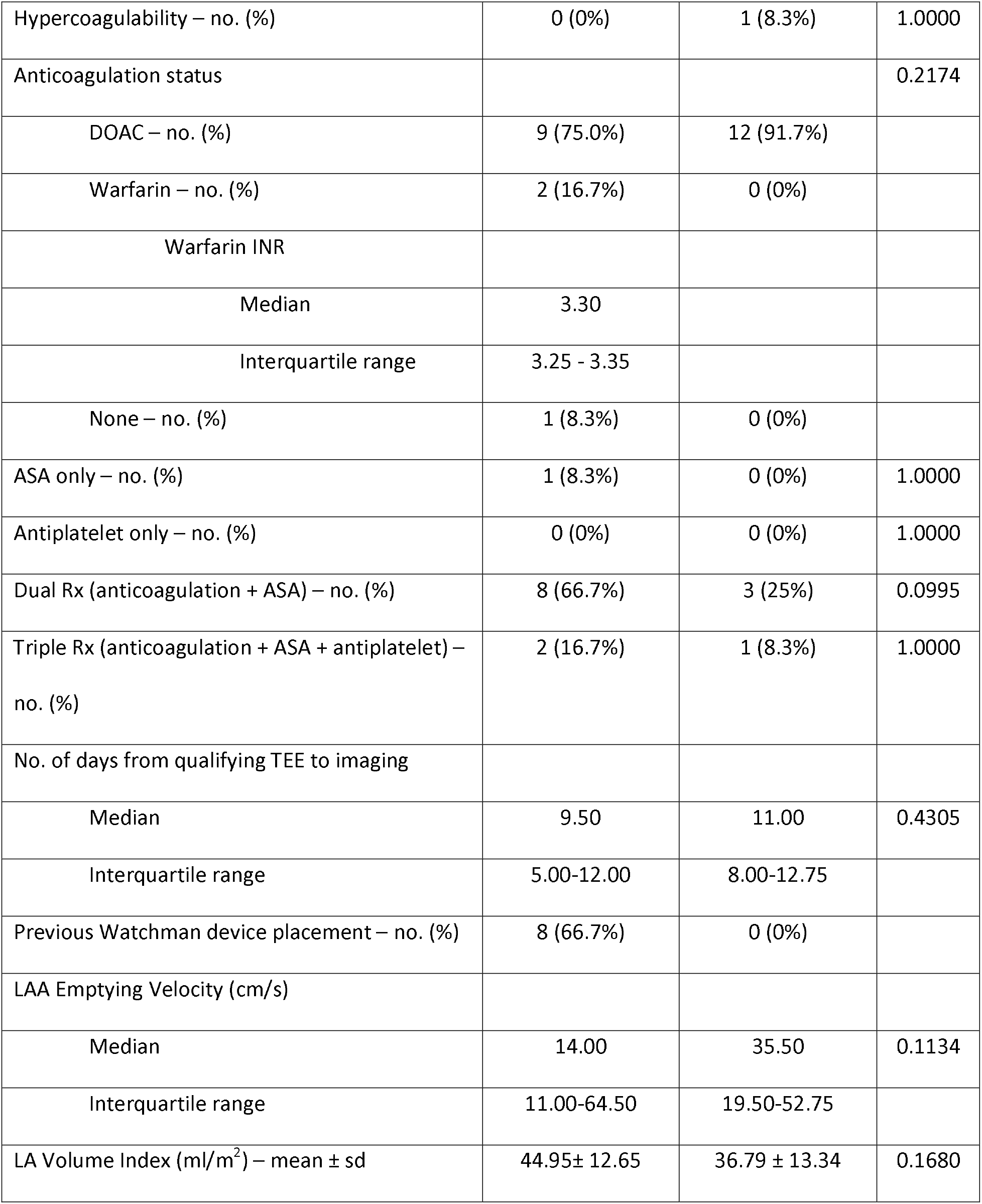
Characteristics of TEE Positive and TEE Negative Subjects

| Characteristic | TEE Positive<br>Patients (N = 12) | TEE Negative<br>Patients (N = 12) | p-value |
| --- | --- | --- | --- |
| Age – yr |  |  |  |
| Median | 80 | 61 | **0.0035** |
| Interquartile range | 76-84 | 55-73 |  |
| Female sex – no. (%) | 4 (33.3%) | 4 (33.3%) | 1.0000 |
| CHA<sub>2</sub>DS<sub>2</sub>-VASc score |  |  |  |
| Median | 4.50 | 2.00 | **0.0101** |
| Interquartile range | 4.00-5.75 | 0.25-3.75 |  |
| GFR (ml/min) – mean ± sd | 54.08 ± 14.23 | 65.55 ± 21.55 | 0.1542 |
| Duration of atrial fibrillation - yr |  |  |  |
| Median | 1.33 | 0.62 | 0.1748 |
| Interquartile range | 0.63 - 2.74 | 0.07 - 1.32 |  |
| Type of atrial fibrillation |  |  | 0.0996 |
| Paroxysmal– no. (%) | 3 (25.0%) | 8 (66.7%) |  |
| Persistent/Permanent– no. (%) | 8 (66.7%) | 4 (33.3%) |  |
| Unspecified– no. (%) | 1 (8.3%) | 0 (0%) |  |
| Current smoking status – no. (%) | 0 (0%) | 1 (8.3%) | 1.0000 |
| Concurrent diagnosis of hypertension – no. (%) | 11 (91.7%) | 8 (66.7%) | 0.3168 |
| Concurrent diagnosis of diabetes – no. (%) | 5 (41.7%) | 0 (0%) | **0.0373** |
| Prior stroke, TIA, or thromboembolism – no. (%) | 5 (41.7%) | 1 (8.3%) | 0.1550 |
| Hypercoagulability – no. (%) | 0 (0%) | 1 (8.3%) | 1.0000 |
| Anticoagulation status |  |  | 0.2174 |
| DOAC – no. (%) | 9 (75.0%) | 12 (91.7%) |  |
| Warfarin – no. (%) | 2 (16.7%) | 0 (0%) |  |
| Warfarin INR |  |  |  |
| Median | 3.30 |  |  |
| Interquartile range | 3.25 - 3.35 |  |  |
| None – no. (%) | 1 (8.3%) | 0 (0%) |  |
| ASA only – no. (%) | 1 (8.3%) | 0 (0%) | 1.0000 |
| Antiplatelet only – no. (%) | 0 (0%) | 0 (0%) | 1.0000 |
| Dual Rx (anticoagulation + ASA) – no. (%) | 8 (66.7%) | 3 (25%) | 0.0995 |
| Triple Rx (anticoagulation + ASA + antiplatelet) – no. (%) | 2 (16.7%) | 1 (8.3%) | 1.0000 |
| No. of days from qualifying TEE to imaging |  |  |  |
| Median | 9.50 | 11.00 | 0.4305 |
| Interquartile range | 5.00-12.00 | 8.00-12.75 |  |
| Previous Watchman device placement – no. (%) | 8 (66.7%) | 0 (0%) |  |
| LAA Emptying Velocity (cm/s) |  |  |  |
| Median | 14.00 | 35.50 | 0.1134 |
| Interquartile range | 11.00-64.50 | 19.50-52.75 |  |
| LA Volume Index (ml/m<sup>2</sup>) – mean ± sd | 44.95± 12.65 | 36.79 ± 13.34 | 0.1680 |

The [^64^Cu]FBP8 PET-MR images in the TEE positive patients typically showed a region of high focal uptake in the LAA compared to the PET signal in the rest of the blood pool and the surrounding tissue. This is shown in Figure 2A-F, where several spontaneous thrombi are present in the LAA by TEE. PET imaging with [^64^Cu]FBP8 detected all of these thrombi. Foci of intense [^64^Cu]FBP8 uptake were detected in the LAA of all the TEE positive patients. In contrast, the majority of TEE negative subjects showed no evidence of focal [^64^Cu]FBP8 uptake in the LAA. This is well demonstrated in Figure 2G-I in a subject with spontaneous echo contrast (SEC) but no thrombus within the LAA. The maximum standardized uptake values (SUV_MAX_) in the LAA were significantly higher in the TEE positive than TEE negative group (median and interquartile range of 4.0 [3.0–6.0] vs. 2.3 [2.1–2.5]; p < 0.001). A SUV_MAX_ threshold of 2.6 resulted in the correct classification of all (12/12) TEE positive subjects and 10/12 TEE negative subjects (Figure 2J-K). This threshold thus produced a positive predictive value of 100% and a negative predictive value of 86%. Receiver operating characteristic (ROC) analysis of SUV_MAX_ produced an area-under-the-ROC-curve (AUROC) of 0.97 (Figure 2L).

**Figure 2.**
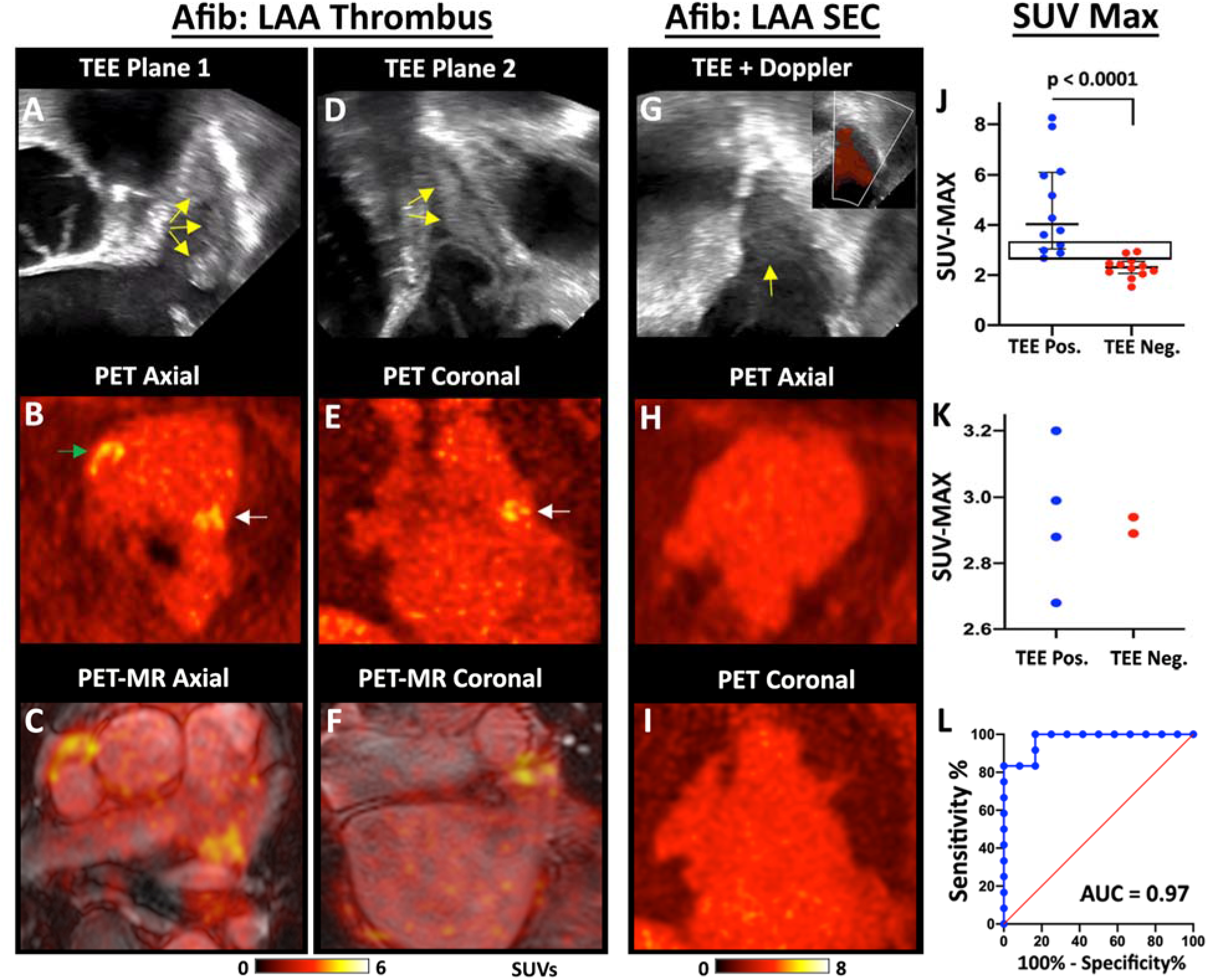
Detection of spontaneous thrombi in the LAA. TEE, PET and fused PETMR images of a subject with multiple LAA thrombi are shown in two orthogonal planes, (A-C) and (D-F). The thrombi produce echogenic/bright foci (yellow arrows) on the TEE images (A, D). The PET (B, E) and PET-MR (C, F) images reveal multiple foci of [^64^Cu]FBP8 uptake in the LAA (white arrows) in both planes. In addition, thrombus is also identified in the RAA (green arrow). (G-I) [^64^Cu]FBP8 does not accumulate in the LAA in a subject with spontaneous echo contrast (SEC) but no thrombus. (G) Echogenic signal is seen within the LAA (yellow arrow), but color Doppler (inset) reveals extensive flow consistent with SEC. No evidence of LAA thrombus is seen on the [^64^Cu]FBP8 PET images in the axial (H) or coronal (I) views. (J-L) The maximum SUV value in the LAA (SUV-Max) accurately distinguishes TEE positive and negative subjects. (J) SUV_MAX_ in the TEE positive subjects was significantly higher than the TEE negative subjects. (J-K) A SUV_MAX_ threshold of 2.6 correctly classified all (12/12) TEE positive subjects and 10/12 TEE negative subjects. The area in the rectangle in panel J is magnified in panel K and confirms that a SUV_MAX_ threshold of 2.6 produces no false negative and only 2 false positive cases. (L) Receiver operating characteristic (ROC) curve demonstrates the robustness of [^64^Cu]FBP8 SUV_MAX_ in distinguishing TEE positive and negative subjects, with an area under the curve (AUC) of 0.97.

The LAA thrombi induced by the placement of the occlusion device (Watchman, Boston Scientific) were extremely conspicuous (Figure 3), likely reflecting the size and high fibrin content in these thrombi. The majority of the Watchman subjects (7/8) were imaged 5–12 days after the procedure. The other Watchman subject was imaged at day 54 and the thrombus in the LAA remained extremely conspicuous (SUV_Max_ of 7.9). All thrombi in the Watchman subjects were confined to the LAA, behind the device, and no evidence of communication between the LAA and left atrium was seen on the bSSFP (balanced steady state free precession) cine images.

**Figure 3.**
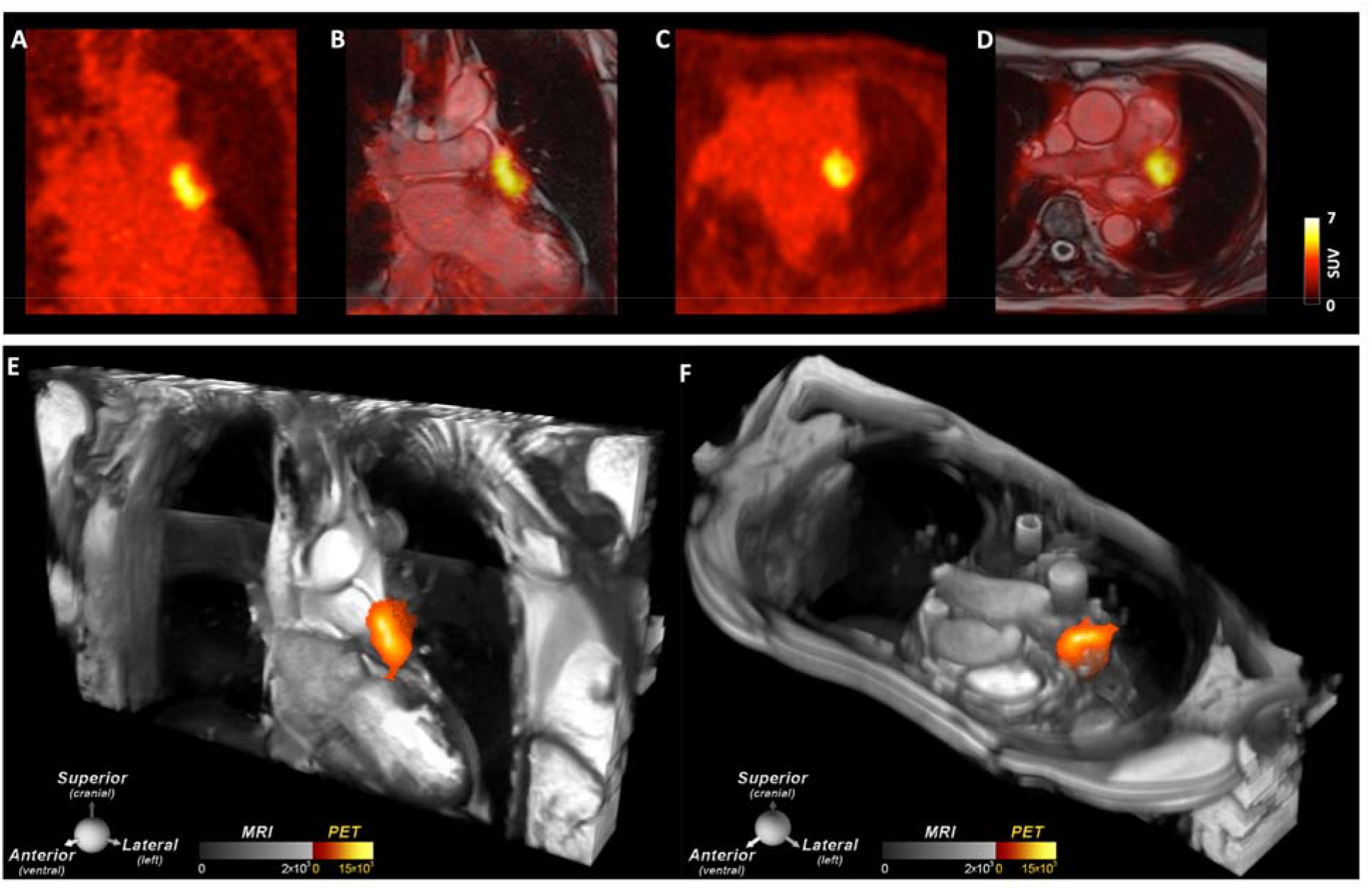
Thrombus in the LAA induced by recent placement of a Watchman LAA closure device. A large thrombus is seen in the LAA behind the device on (A, B) the coronal PET and PET-MR images as well as (C, D) the axial PET and PET-MR images. (E, F) 2D bSSFP cines stacks in the coronal and axial planes have been combined into a single 3D dataset and fused with the 3D PET data. In these multi-modal images the PET and MR data are both displayed in 3D, creating a volumetric depiction of the heart and the thrombus/[^64^Cu]FBP8 containing LAA. Volume rendered images in the oblique coronal and axial planes confirm the presence of thrombus in the LAA. (A-F) The thrombus produced by the closure device is contained within the LAA, and no evidence of thrombus is seen elsewhere in the heart or thorax.

### Integrated Approach with [^64^Cu]FBP8 PET and MR T1 Mapping

The degradation of red blood cells trapped within a thrombus can result in the production of paramagnetic methemoglobin, which shortens the longitudinal relaxation time (T1) of blood. We, therefore, next investigated the value of T1 mapping of the LAA, particularly in combination with PET imaging of [^64^Cu]FBP8. T1 mapping was performed with a Modified Look Locker Imaging sequence, which only became available on our PET-MR scanner after the first 6 subjects had been imaged. Consequently, T1 mapping was performed in all the TEE positive subjects in the study but only 6 of the TEE negative subjects. Inspection of the individual T1 weighted images revealed the presence of hyperintense signal in the LAA (Figure 4A, 4E), consistent with LAA thrombus in this TEE positive subject. T1 mapping confirmed the presence of large foci of reduced T1 in the LAA, which were extremely conspicuous in the Watchman subjects (Figure 4B-C). Fusion of the PET images and T1 maps showed a high degree of colocalization between areas with high SUVs and reduced T1 (Figure 4D). The detection of spontaneous LAA thrombi with T1 mapping was more challenging, likely due to their smaller size and more advanced age, but still highly feasible (Figure 4E-H). Fusion of the T1 maps with Dixon fat and water images was extremely helpful in ensuring that the foci of short T1 were not regions of fat, but rather areas of water rich thrombus.

**Figure 4.**
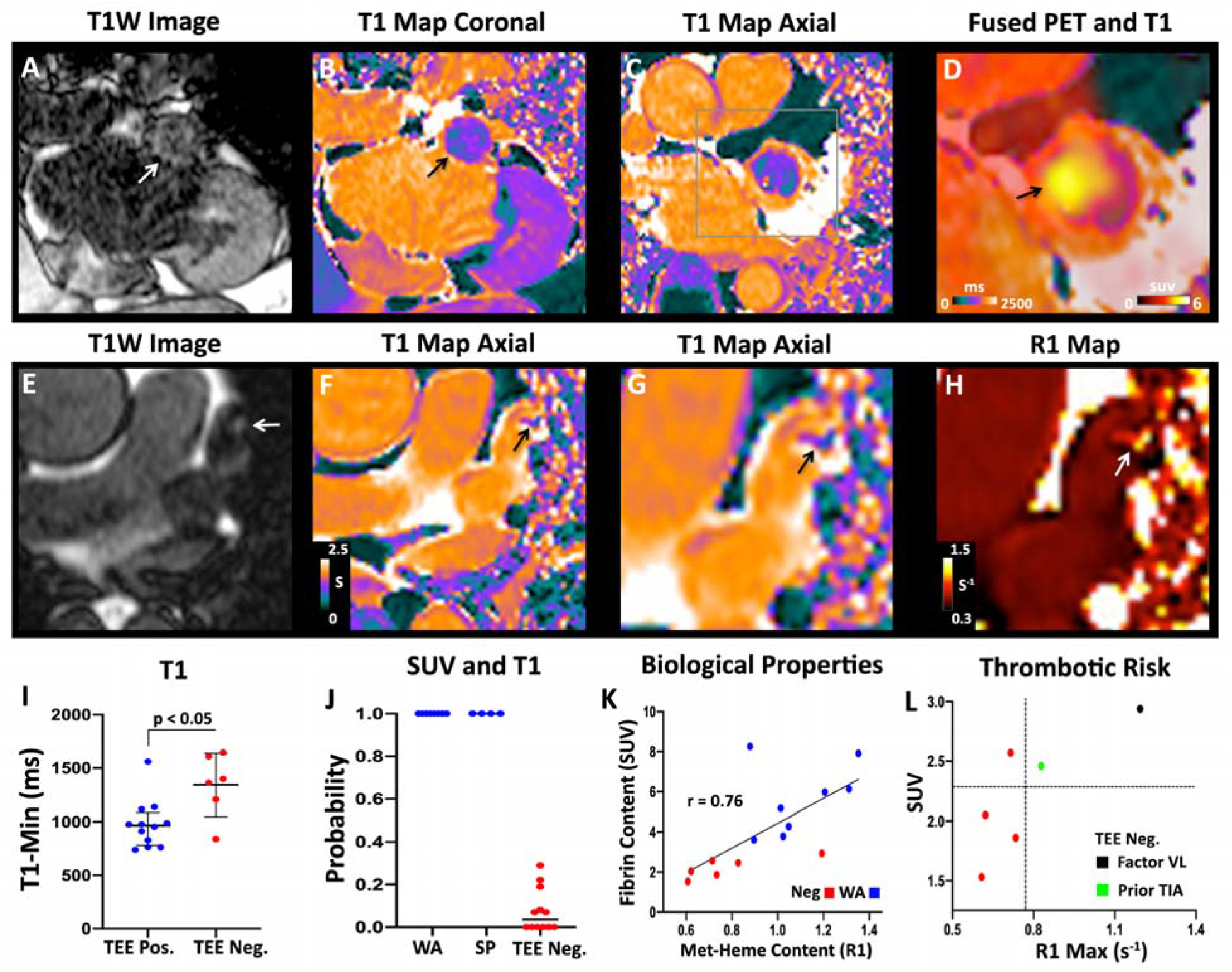
Integrated analysis of magnetic relaxation (T1) and SUV_MAX_ in the LAA. (A-D) Subject with a LAA closure device. (A) T1W image in the coronal plane showing a hyperintense region in the LAA (white arrow). (B) T1 map in the same view demonstrating low values (green-purple; black arrow) in the LAA, similar to the T1 of myocardium (purple) but far lower than the blood pool (orange). (C) Axial T1 map confirms the presence of short-T1 (paramagnetic) species in the LAA, consistent with methemoglobin (met-heme) accumulation in thrombus. (D) Magnified view of the axial T1map fused with the [^64^Cu]FBP8 PET image demonstrates a high degree of the overlap between low T1 and high [^64^Cu]FBP8 uptake. (E-H) Spontaneous LAA thrombus. (E) Axial T1W image showing a hypertintense focus (arrow) in the LAA. (F) Axial T1 map shows a focus of reduced T1 (arrow) in the LAA. (G) Magnified view of T1 map and short T1 focus (arrow). (H) Magnified view of R1 map (units s^−1^). The thrombus in the LAA (arrow) has a high R1 value. (I) The minimum T1 value (T1_MIN_) in the LAA is significantly shorter in the TEE positive than negative group. (J) Probability of having a LAA thrombus based on logistic regression of the z-scores for both SUV_MAX_ and T1_MIN_. WA = Watchman, SP = spontaneous. (K) Biological properties of the LAA (fibrin and met-heme content) in those with precisely aged recent thrombi (WA group) and no thrombi (Neg group). A strong correlation is seen between fibrin content (SUV_MAX_) and met-heme content (R1_MAX_ = 1/T1_MIN_). (L) Within the TEE negative group an association may be present between higher SUV and R1 values and thrombotic risk. Factor VL = Factor V Leiden.

Quantitative analysis of the T1 maps was performed by measuring the minimum T1 value (T1_MIN_) in the LAA, analogous to SUV_MAX_ in the PET images. As shown in Figure 4I, T1_MIN_ was significantly shorter in the TEE positive subjects compared to the TEE negative subjects 970 [780–1080] versus 1380 [1120–1620], respectively, p < 0.05; although there was some overlap in the T1_MIN_ values between groups. Joint analysis of the SUV and T1 data was then performed using logistic regression. The z-scores for SUV_MAX_ and T1_MIN_ were used as inputs. For each SUV_MAX_ or T1_MIN_ value, the corresponding z-score describes the number of standard deviations it is above/below the mean value in the blood pool (left ventricle for the PET images, and left atrium or right pulmonary artery for the T1 maps). Logistic regression using the combined zscores of SUV_MAX_ and T1_MIN_ was extremely accurate (Figure 4J). All TEE positive cases were detected with a probability of 100% and no TEE negative case had a probability > 30%. The diagnostic accuracy of this approach was 100%, with an AUROC of 1.

We next examined the relationship between fibrin and methemoglobin (met-heme) content in the LAA, measured by SUV_MAX_ and R1_MAX_, respectively, where R1_MAX_ = 1/T1_MIN_. The correlation between these measures in spontaneous LAA thrombi was poor (see Supplement), perhaps reflecting the heterogeneous nature of this group. However, in the remaining 14 subjects in whom both SUV and R1 data were available(8 Watchman and 6 TEE negative) a strong correlation (r = 0.76) was seen between fibrin and met-heme content in the LAA (Figure 4K). Further, in the 6 TEE negative subjects, the presence of both high SUV_MAX_ and R1_MAX_ values in the LAA was associated with high risk features, such as a Factor V Leiden mutation and a history of cerebral thromboembolism (Figure 4L).

### Composite Detection of Cardiac and Extracardiac Thrombi

Qualitative analysis of the data for the presence of cardiac and extracardiac thrombi was performed by a radiologist blinded to the TEE data. While quantitative analysis with the SUV_MAX_ threshold described above produced a sensitivity of 100%, qualitative inspection of the images correctly classified 11/12 TEE positive subjects and 12/12 TEE negative subjects. Co-incidental thrombi were relatively uncommon and included 1 pulmonary embolism (Figure 5A-C) and right atrial appendage thrombus in 4/24 subjects. One of the subjects in the study had a history of a large intracranial bleed, which was approximately 9 months old at the time of imaging (Figure 5D-F). Only faint signal was seen on the perimeter of the site of the bleed, suggesting that this old injury now comprised organized thrombus with low fibrin content.

**Figure 5.**
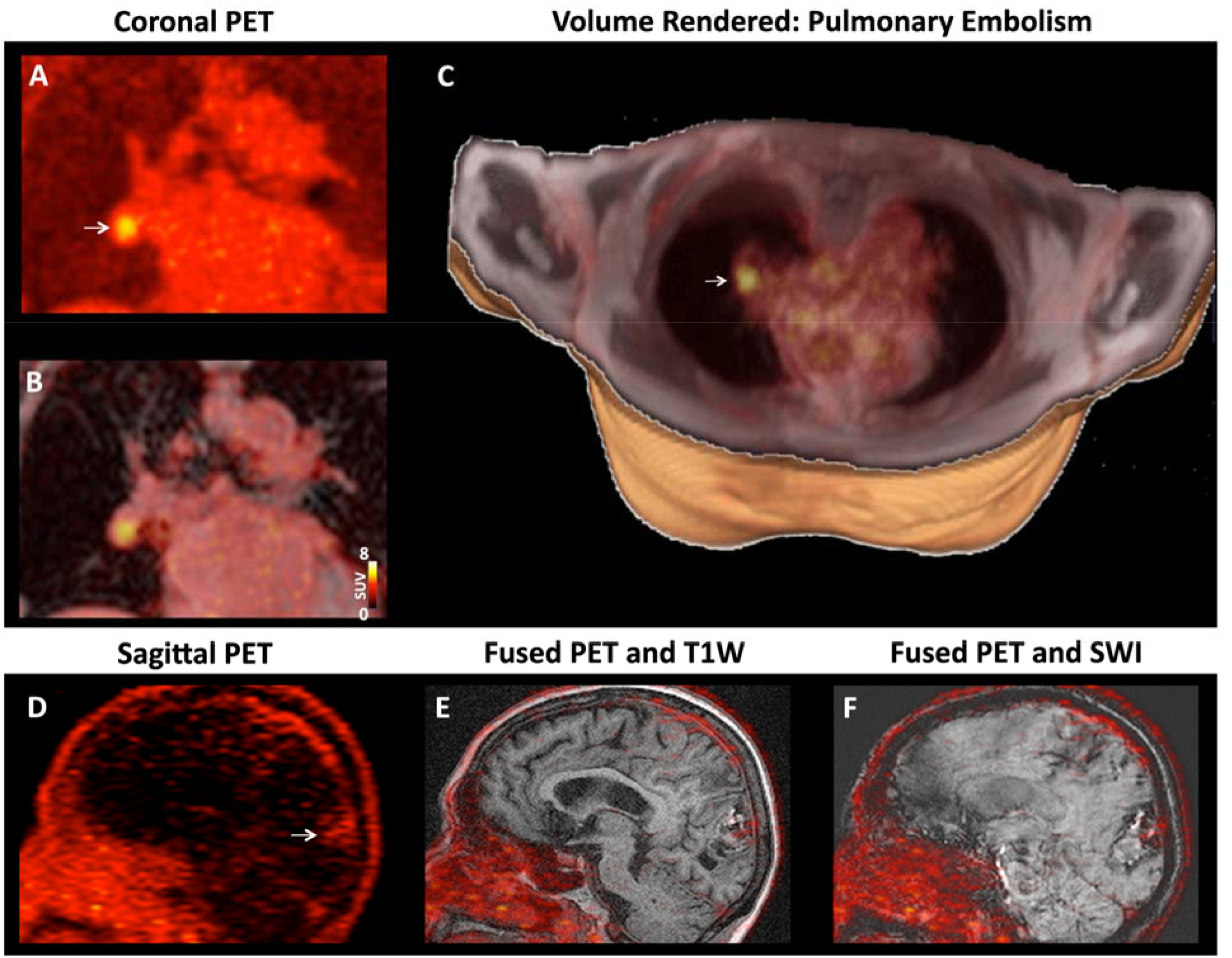
[^64^Cu]FBP8 facilitates whole-body thrombus detection. (A-C) Right pulmonary artery embolism in a subject with a spontaneous LAA thrombus. (A) Coronal PET image with a focus of high [^64^Cu]FBP8 activity (arrow). (B) Fusion of the PET and Dixon water images reveals that the thrombus is located within a branch of the right pulmonary artery. (C) Volume rendered image showing the pulmonary embolism (arrow,) in the right lung. (D-F) TEE positive subject with a history of a large intracranial bleed. (D) Sagittal PET image, (E) fused PET and T1-weigted image and, (F) fused PET and susceptibility weighted image (SWI). The PET image shows that a low level of [^64^Cu]FBP8 uptake persists at the site of the bleed (arrow).

## Discussion

Anticoagulation is recommended in many clinical scenarios but is frequently empiric and complicated by the risk of major, and potentially fatal, hemorrhage (*1, 2*). Here, we present a translational molecular imaging approach, based on the detection of fibrin in thrombi, with the potential to transform the diagnosis and therapy of thrombosis. We show in this first-in-human study that PET-MR of the fibrin-binding PET probe [^64^Cu]FBP8 can detect acute to subacute LAA thrombi with a sensitivity of 100% and specificity of 84%, matching the performance of current more invasive approaches. The probe allows the composite burden of thrombosis throughout the body to be imaged and provides a specific molecular signature that directly reflects the biological properties of thrombi.

Human thrombus molecular imaging has been attempted in the past with limited success (*3*). For instance,^99m^Tc-apcitide utilized an RGD-based peptide targeting the glycoproteinII_b_/III_a_ (GP2B3A) receptor on activated platelets (*7, 8*). While approved by FDA for DVT imaging in 1998, it was not widely used due to poor efficacy. Multicenter Phase III data showed sensitivity and specificity for DVT of only 73% and 68% compared to a gold standard of contrast enhanced venography (*9*).^99m^Tc-apcitide was also shown to be ineffectual for diagnosis of PE (*5*). More recently, a PET probe targeting the GP2B3A receptor has been used to successfully image acute arterial and venous thrombi (*4, 6*). However, the specificity of this agent remains unknown, and the targeting of activated platelets, through the GP2B3A receptor, supports the detection only of active, hyperacute platelet-rich thrombi. Immunological approaches to fibrin imaging with radiolabeled antibodies or antibody fragments have been reported for diagnosis of deep vein thrombosis (*12–16, 20)*, but these were all abandoned because of low sensitivity and specificity (*21*). More recent work has involved an antibody (*19, 20*), and antibody fragment (*17, 18*), to the D-dimer region of cross-linked fibrin, which remains accessible even after thrombus formation has been stopped by anticoagulant treatment. While targeting the D-dimer region has improved detection, these agents are still limited by a long plasma half-life, i.e. 21 hrs for the antibody (*20*) and 12 hrs for the antibody fragment (*18*)), resulting in high blood background or the need for very delayed imaging, e.g. next day.

The development and application of [^64^Cu]FBP8 was based on the rational design of its preclinical precursors (*27–30*). The fibrin-targeting peptide on the probe was obtained from phage display screens via negative selection against fibrinogen and serum albumin, followed by positive selection against fibrin to engineer specificity (*25*). This resulted in high affinity for fibrin, but very low non-specific binding to fibrinogen or other plasma proteins. A similar peptide was previously conjugated to 4 gadolinium chelates to form a MR-detectable agent, EP-2104R (*11, 36*), which was evaluated in humans in Phase 2a proof-of-concept studies (*26, 37*). Thrombus enhancement was impressive, but these studies demonstrated that imaging was best performed ≥ 2 h post injection. For MRI, this necessitates two separate imaging sessions (pre and post probe injection) and is a major limitation. While no cases of nephrogenic systemic fibrosis have been reported with EP-2104R, the use of gadolinium-based probes in subjects with renal impairment remains a concern. A strong case existed, therefore, to develop a PET-based probe based on this peptide. A medicinal chemistry effort ensued to construct a peptide-chelate conjugate with optimized affinity for fibrin, prolonged metabolic stability, and rapid plasma pharmacokinetics (*27, 28, 38–41*). [^64^Cu]FBP8 emerged from this process with nanomolar affinity to fibrin, rapid blood clearance and absence of metabolism in rodents, and high sustained and specific uptake in different animal models of arterial, venous, and embolic thrombi (*28–30*). [^64^Cu]FBP8 performed similarly in the human subjects studied here to preclinical studies with the probe, with its relatively rapid blood clearance allowing imaging to be performed 1 hour after injection, thus simplifying the logistics of clinical translation.

Many targets that have been explored for thrombus detection are highly expressed in acute but not chronic thrombi. While the level of fibrin in thrombi does decrease with time (*30, 42*), our data suggest that it remains sufficient to support detection with [^64^Cu]FBP8 for at least 8–10 weeks. The age of spontaneous LAA thrombi is difficult to discern, however, we estimate based on serial TEEs that one of the spontaneous thrombi detected in the study may have been up to 9 weeks old. In addition, one of the subjects with a Watchman device was imaged 54 days after implantation. The SUVs in the thrombi over this range (< 10 weeks) remained stable. Further study will be needed to determine the sensitivity of [^64^Cu]FBP8 for chronic thrombi (> 10 weeks), the utility of adding a gadolinium-based contrast agent to facilitate their detection, and the embolic risk of these fibrin-poor organized thrombi.

The degradation of red blood cells in subacute thrombi results in the production of the strongly paramagnetic met-heme species, which can be detected with T1 mapping (*43*). This approach requires no exogenous contrast and also exploits the superior spatial resolution of MRI. When integrated with PET imaging of [^64^Cu]FBP8, the combined approach detected the presence or absence or thrombus with 100% accuracy. In addition, rather than merely detecting a filling defect, relevant biological information on the properties of the thrombus could be obtained (*43*). In large acute to subacute thrombi (Watchman subjects), and interestingly in the TEE negative subjects as well, a strong correlation was seen between fibrin and met-heme content in the LAA. High SUV_MAX_ and R1_MAX_ values in the TEE negative subjects were also associated with high risk features such as the presence of a hypercoagulable state (Factor V Leiden) or history of cerebral thromboembolism. While very preliminary, these observations in the TEE negative cohort raise the possibility that small microthrombi, undetectable by TEE, may be present in the LAA during atrial fibrillation (*44–46*). Studies using transorbital Doppler in TEE negative subjects have documented multiple embolic signals to the brain, supporting the presence of subclinical microthrombi in atrial fibrillation. In subjects with spontaneous LAA thrombi the correlation between fibrin and met-heme content in the thrombus was poor. This likely represents the heterogeneous nature of this cohort and the older age of their thrombi. Collectively, while very preliminary, the observations above demonstrate the potential value of integrated PET-MR imaging with [^64^Cu]FBP8, T1 mapping and other forms of endogenous MR contrast (magnetization transfer, diffusion) with sensitivity to thrombi (*47*), and underscore the promise of these systems.

Several limitations of this study merit discussion. The PET images of [^64^Cu]FBP8 were not gated or motion corrected resulting in volume averaging of the [^64^Cu]FBP8 PET signal in the LAA, and which likely results in underestimation of probe uptake in the thrombus. While PET-MR avoided the additional radiation exposure inherent in PET-CT, and allowed us to perform T1 mapping studies to provide added diagnostic power and molecular characterization, attenuation correction of thoracic PET-MR data is more complex and still an area of active research (*48*). Although we minimized potential artifacts introduced by the attenuation correction method supplied by the scanner manufacturer, remaining inaccuracies could have biased our measurements. Further work will be needed to develop attenuation and motion compensation strategies tailored to atrial fibrillation, where the irregularity of cardiac cycle length creates a major challenge. The pharmacokinetic studies in healthy volunteers indicated that the blood background signal continued to clear over several hours. However, for logistical reasons, the atrial fibrillation subjects were imaged one hour after [^64^Cu]FBP8 injection, which resulted in higher background signal. Despite these limitations, the LAA thrombi were highly conspicuous with high thrombus to background ratios. The number of spontaneous LAA thrombi in the study was low but sufficient, we believe, to demonstrate proof-of-principle in this first-in-human study. Likewise, while the study did not focus on the detection of thrombi outside the LAA, when these were encountered incidentally, in the RAA and pulmonary artery for instance, they were also highly conspicuous.

In summary, we present the first human experience with a novel fibrin-binding radiotracer, [^64^Cu]FBP8, and show that it facilitates the detection of acute-subacute LAA thrombi with a sensitivity of 100% and a diagnostic accuracy greater than 90%. We also show that the integrated use of [^64^Cu]FBP8 and T1 mapping of the LAA can further increase diagnostic accuracy and characterize aspects of thrombus biology. The probe is well tolerated, demonstrates favorable pharmacokinetics and can support whole body detection of thrombosis. [^64^Cu]FBP8 has the potential to advance the understanding of thrombus biology and to transform the diagnosis and treatment of thrombosis in a wide range of disease settings.

## Materials and Methods

### Synthesis of [^64^Cu]FBP8

The precursor molecule without the Cu-64 is formulated at a concentration of 90 ± 10 μ g/mL in 100 mM sodium acetate buffer, pH 5.5, and kept frozen as 1 mL aliquots in 20 mL vials until needed for radiolabeling. For labeling, the precursor is allowed to warm to room temperature, up to 1.5 GBq [^64^Cu]CuCl_2_ in HCl solution is added, the solution is diluted to 10 mL by addition of saline, and then the vial is heated to 60°C for 40 min to yield [^64^Cu]FBP8, requiring no additional purification.

### Study Subjects

This study was approved by the Partners Institutional Review Board (protocol number 2015P002385) and registered on the http://ClinicalTrials.gov website (NCT03830320). All subjects were aware of the benefits/risks of the study and provided written consent. Healthy controls (HC) were recruited via the Partners Healthcare System Research Study Volunteer Program, and had to be 18 years of age or older with no known medical issues, have no history of atrial fibrillation or thrombosis, and a negative drug screen. Patients with atrial fibrillation (Afib), presenting to the electrophysiology laboratory at the Massachusetts General Hospital (MGH) for TEE guided cardioversion, were recruited for the LAA arm of the study. These subjects fell into three groups: those with no sign of LAA thrombus in the TEE (n = 12), those with spontaneous thrombi in the LAA on TEE (n = 4), and those in whom thrombus in the LAA was purposefully induced through the placement of a LAA closure device (Watchman, Boston Scientific, n = 8). Most of the subjects (7/8) with Watchman devices were imaged within 12 days of the device being placed. One Watchman subject was recruited from the echocardiography laboratory at the MGH when presenting for his scheduled TEE 6 weeks after the placement of the device. All subjects underwent PET-MR with [^64^Cu]FBP8 within 14 days of their reference TEE. Subjects were excluded if they had electrical implants such as a cardiac pacemaker/defibrillator, exceeded the weight limit of the PET-MR scanner or had a contraindication to MRI. Further details on the clinical characteristics of the TEE negative and TEE positive study subjects are provided in Table 1.

### PET-MR Data Acquisition

All images (HC and AFib) were acquired on a simultaneous PET/MR scanner (Biograph mMR, Siemens Heathineers, USA).

### Healthy Controls (HC)

For the HC subjects multi-bed whole-body PET images were continuously acquired (4 min per bed position) in 3D listmode starting from the injection of an average dose of 380 (full range 210 – 530) MBq of [^64^Cu]FBP8 for up to 2 h post injection (p.i.), the subject was returned to the scanner for a second scan at 4 h p.i. and, if they agreed, the next day for a third scan. In order to have similar reconstructed images in all subjects, a single static image was retrospectively reconstructed in all healthy subjects around ∼70 min p.i.. PET images were reconstructed using a 3D ordinary Poisson ordered subset expectation-maximization (OP-OSEM) algorithm (*49*) with 3 iterations and 21 subsets, using corrections for attenuation, scatter, randoms, delays, dead-time, background, sensitivity, normalization and radioisotope-decay.

Simultaneous to the PET acquisition, MRI of the thorax was performed using the product 6-channel Body matrix surface coil of the scanner. The following MR images were acquired for the HC: a dual-echo Dixon-VIBE sequence (TR = 3.96 ms; TE = 1.23/2.46 ms; flip angle = 9º, reconstruction matrix = 192×120; voxel size = 2.6×2.6×3.1 mm^3^; total acquisition time = 6sec); and a T2 Haste sequence, acquired coronally (TR = 1000 ms; TE = 100 ms; TI = 200 ms; flip angle = 120º, reconstruction matrix = 256×256 voxel size = 1.6×1.6×6.9 mm^3^; total acquisition time ∼30sec). Both these sequences were acquired during breath-hold at end expiration. The attenuation maps generated from the Dixon-VIBE images using the standard method provided by the manufacturer were inspected for all the subjects. Noticeable artifacts (e.g. lung and soft tissue misclassification) were corrected using a semiautomatic procedure by an image processing expert blinded to the results of the [^64^Cu]FBP8 data analysis (CC).

### Afib Subjects

For the AFib subjects PET images of the thorax were acquired for 45 min (AFib) in 3D listmode starting at 70 min p.i. of and average dose of 250 (full range 120 – 450) MBq of [^64^Cu]FBP8 radiotracer. PET images were reconstructed using the same method and parameters as for the HC. In those subjects who were agreeable, additional imaging was performed of the neck and head (7.5 minutes at each station). The majority (20/24) of Afib subjects were in sinus rhythm at the time of imaging. No cardiac or respiratory gating was used in the PET reconstructions in any of the subjects.

Simultaneous to the PET acquisition, the dual-echo Dixon-VIBE and HASTE sequences were acquired as described above. The attenuation maps produced by the scanner were inspected and corrected, if necessary, as described above. In addition, single shot bSSFP images were acquired in the axial plane to identify the LAA. ECG-gated bSSFP cine images were acquired through the LAA in the axial and coronal planes using the following parameters: TR = 51.15 ms; TE = 1.5 ms; flip angle = 50º; reconstruction matrix = 208×256; voxel size = 1.41×1.41×7 mm^3^ and 25 cardiac phases; Maps of native T1 in the LAA were acquired in the same axial and coronal planes using the product Modified Look-Locker imaging (MOLLI) sequence with the following parameters: TR = 281 ms TE = 1.12 ms; TI = 100–3090 ms; flip angle = 35º, reconstruction matrix = 256×218; voxel size = 1.4×1.4×8 mm^3^). The MOLLI sequence selected was determined by the patient’s heart rate (< or > 85 bpm) and, if off-resonance artifacts were present, frequency scouting was performed to select the optimal center frequency. The MOLLI sequence was installed on our PET-MR scanner after the first 6 TEE negative subjects had already been scanned. T1 mapping was, thus, performed in 6/12 TEE negative subjects and 12/12 TEE positive ones.

### Analysis of Blood Samples

For the healthy volunteers, blood samples were drawn from a venous catheter before and after intravenous injection of [^64^Cu]FBP8. A venous line was placed in the arm opposite the site of injection. This second IV line was used to draw up to 10 blood samples of 6 mL volume in EDTA collection tubes. The line was flushed with saline between blood draws.

The plasma was separated, weighed, and counted on a gamma counter. The activity in the blood at each time point was computed as percentage of injected dose per gram (%ID/g) by dividing the measured activity per gram by the activity in the injected dose. The %ID/g versus time data was fit to an exponential function to estimate plasma half-life.

To assess for metabolites, an aliquot of plasma was mixed 1:1 with ice cold acetonitrile (ACN) and solids were separated by centrifugation. The supernatant was injected onto an analytical HPLC (Agilent 1100) with a C18 reverse phase column, and eluted with a gradient of 5% ACN: 95% water, each containing 0.1% trifluoroacetic acid (TFA), to 95% ACN/TFA over 15 minutes. The eluent was collected every 30 s, and the activity of each fraction was measured using a gamma counter. The result was compared with intact probe spiked into plasma from the subject taken before injection.

To determine if the activity in the plasma was capable of binding to fibrin, a functional assay was performed (*36*). Briefly 100 µL of human fibrinogen was aliquoted into a 96-well plate at a concentration of 2.5 mg/mL, then thrombin (1 U/mL) and 7 mM CaCl_2_ were added to polymerize to fibrin, and the plates were dried to a thin film overnight at 37 °C. For the functional assay, 100 µL of plasma were added to wells containing fibrin and to blank wells and incubated on a rocker for 2 h. Aliquots of the plasma were removed and counted and the fraction bound was calculated as the (activity from the blank well – activity in fibrin well)/(activity in blank well). The fraction bound at each time point was then compared to the fraction bound measured when pure [^64^Cu]FBP8 was spiked into plasma from the subject taken before injection.

### PET Image Analysis

Signal was measured in two regions of interest (ROIs) in the PET images: The LAA and the left ventricular blood pool. To avoid bias, the ROIs were drawn using OsiriX MD (v.10.3, Pixmeo, Switzerland) on the T2 Haste images (HC) or the Dixon opposite phase images (Afib subjects) and then copied without modification to the PET images. A large spherical ROI was drawn to enclose the entire LAA and was standardized as much as possible across the subjects.

PET images were converted into standardized uptake values (SUV) using the subject’s weight as a normalization factor (*50*). Mean and maximum SUVs (SUV and SUV_max_ respectively) were obtained from the ROIs after exporting the OsiriX ROIs into Matlab for data analysis. ROC analysis of the SUV_MAX_ values was performed in GraphPad Prism and a SUV_MAX_ threshold of 2.6 was chosen from the ROC curve to provide 100% sensitivity.

### T1 Map Analysis

T1 maps were produced on the scanner using the reconstruction package provided by the manufacturer (Myomaps, Siemens Healthineers). Three ROIs were drawn using OsiriX in each coronal and axial T1 map: *LAA*, a manually drawn ROI that followed closely the contour of the anatomical region of the LAA; *BloodPool_T1_*, a manually drawn ROI on either the left atrium or the right pulmonary artery; and *Fat*, a subcutaneous fat ROI near the LAA. ROIs were exported into an in-house Matlab program and mean and minimum T1 values (T1 and T1_min_ respectively) were obtained from the LAA. To prevent voxels within the adipose tissue surrounding the LAA from biasing the analysis, voxels within the LAA with values lower than two standard deviations above the mean of the

*Fat* region were excluded.

### Combined PET-MR Analysis: Z-Score Analysis with Logistic Regression

Z-scores are defined as the number of standard deviations above (for PET images) or below (for T1 maps) the mean of the background region. For PET data the SUV_max_ in the LAA and mean SUV in the blood pool are used to derive the z-score. Likewise, for T1 mapping, the T1_MIN_ in the LAA and mean T1 in the blood pool are used. The z-score is defined mathematically as:

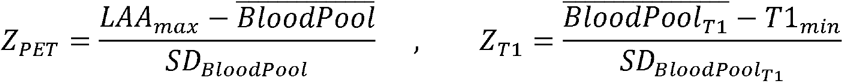

Z-scores define a normalized parameter that helps minimize the potential impact of variability across subjects, such as variable PET uptake due to different biodistribution and/or pharmacokinetic patterns (for PET images) or inter-scan variability (both PET and T1 maps).

Z-scores for PET and T1 were used in a logistic regression analysis to estimate the predictive value of combined PET and MR measurements to define a subject *j* as positive, with respect to the gold standard of TEE, as follows:

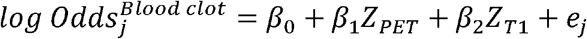

where β_0_ is the intercept of the model, β_1_ and β_2_ are the log of the odds ratio for the Z_PET_ and Z_T1_ respectively, and *e_j_* is the error of the estimation for subject *j*.

### Qualitative Analysis

Qualitative analysis of the PET images was performed by a radiologist with more than 15y of experience (O.C.), who was blinded to the TEE data.

### Statistical Analysis

Descriptive statistics were calculated to summarize clinical characteristics. Differences between the TEE positive and negative groups were assessed using a two-tailed Student’s t-test for normally distributed characteristics, and a Mann-Whitney test for non-normally distributed characteristics. A Fisher’s exact test (with Freeman-Halton extension for multiple comparisons) was used for dichotomous outcomes. All datasets were tested for normality (GraphPad Prism) using the D’Agnostino-Pearson omnibus K2 test. The maximum SUV value (SUV_MAX_) in the LAA of the TEE positive and negative subjects was compared using a Mann-Whitney test (GraphPad Prism). The minimum T1 value (T1_MIN_) in the LAA of the TEE positive and negative subjects was also compared using a Mann-Whitney test (GraphPad Prism). Median values with interquartile ranges are reported for both SUV_MAX_ and T1_MIN_. Correlations were calculated using the Spearman coefficient. Logistic regression was used, as explained above, to estimate the predictive value of the combined PET-MR measurements. All statistical tests were run on Graphpad Prism v.8.4.0 or Rstudio v.1.2 using R package v.3.6.2.

## Data Availability

All data associated with this study are available in the main text. All data can be obtained from the authors under reasonable request.

## Acknowledgments

We thank the members of the radiopharmacy laboratory at the Martinos Center for their assistance in the preparation of [64Cu]FBP8. We thank the nurses and physicans in the electrophysiology laboratory at the MGH for their co-operation in subject recruitment. We thank the PET-MR technologists in the Martinos Center (Grae Arabaz, Shirley HSU and Regan Butterfield) for their assistance.

